# Study Protocol for a Randomized Controlled Trial Comparing Swiss Ball Balance Training and Conventional Balance Training to Improve Trunk Control and Mobility in Individuals with Sub acute Stroke

**DOI:** 10.64898/2026.09.11.26362878

**Authors:** Brian Nkhoma, Miriam Mapulanga

## Abstract

Stroke is a leading cause of mortality and long-term disability worldwide, with low- and middleincome countries carrying the greatest burden. Impaired trunk control and balance are common after stroke, severely limiting mobility and independence and increasing the risk of falls. Conventional balance training is widely used to improve stability, but it primarily targets static postural control and may not sufficiently activate trunk muscles during dynamic tasks. Swiss ball balance training, which uses an unstable surface to engage core musculature and enhance sensorimotor integration, has shown promising results in improving trunk control and functional mobility. However, research comparing Swiss ball balance training with conventional balance training is limited, particularly in Sub-Saharan Africa. This study aims to compare the effects of Swiss ball training and conventional balance training on trunk control, balance, and mobility in individuals with subacute stroke.

A randomized, assessor-blinded, two-period, two-sequence crossover trial will be conducted. Participants will be randomly assigned to Swiss Ball Balance Training or Conventional Balance Training, each administered for 45 minutes per session, three days per week, over six weeks, with a one-week washout period between phases. Standardized clinical outcome measures will assess trunk control, postural stability, and functional mobility at baseline and after each intervention phase. Data will be analyzed using paired t-tests and Wilcoxon rank-sum tests in Stata v17 (p < 0.05).

It is expected that Swiss ball exercises will yield greater improvements in trunk control, balance performance, and functional mobility than conventional methods. The results are expected to help design context-specific rehabilitation programs.

## Background

This study protocol was developed to compare Swiss ball balance training (SBBT) with conventional balance training (CBT) to improve trunk control and mobility in individuals with sub-acute stroke at St. Francis’ Hospital (SFH) in Katete district of Zambia. Stroke is a neurological condition of vascular origin, characterized by impaired motor control and significant limitations in mobility (1). Globally, it is the second leading cause of death among individuals over 60 years and the fifth leading cause of death in those aged 15–59 (2,3,4). Low- and middle-income countries (LMICs) carry a disproportionate burden, accounting for approximately 70% of stroke-related deaths and 87% of stroke-related disabilities (5). In 2021, the global prevalence of stroke was estimated at 93.8 million cases, with 11.9 million new cases reported (6).

Beyond mortality, stroke is a leading cause of long-term disability, with one-third of survivors permanently disabled (2). Stroke-related disabilities encompass sensorimotor, coordination, cognitive, language, and emotional impairments (7). Among the most common deficits are poor trunk control and balance, which are critical for mobility and activities of daily living (ADLs) (1,8). Impaired trunk stability contributes to difficulties in maintaining sitting and standing balance, altered gait, and increased fall risk (9,10). Approximately 83% of stroke survivors experience some degree of balance dysfunction (3,10,11), underscoring the need for targeted rehabilitation strategies (11).

Trunk control precedes limb movement and is closely correlated with functional mobility (9). This is because trunk muscles are essential for maintaining anti-gravity postures and provide a stable foundation for both static and dynamic activities (7). They ensure dynamic stability of the spine and pelvis, enabling effective weight shifts during movement against gravity (12).

Rehabilitation approaches commonly include conventional balance training (CBT) and Swiss ball balance training (SBBT) (13). The Swiss ball, also known as a physio ball or gym ball, is an unstable device widely used in fitness and rehabilitation (14). Exercises performed on a Swiss ball increase trunk muscle demand, promoting greater force generation to maintain stability and balance. In routine practice, CBT typically involves static postural exercises such as reaching and core stability training, selected at the therapist’s discretion (7). While CBT has long been the standard approach for retraining balance and trunk control (14), it may not fully engage trunk muscles under dynamic, unstable conditions that better simulate functional movements(15).

Early integration of effective balance-focused rehabilitation in stroke recovery has been shown to improve outcomes, enhance functional independence, and increase quality of life (16). This highlights the importance of implementing evidence-based rehabilitation strategies that prioritize balance improvement within stroke care guidelines.

### Main Research Objective

To compare the effect of Swiss Ball balance training over conventional balance training on trunk control and mobility performance on individuals with sub-acute stroke

### Specific Research Objectives

To assess the effect of Swiss ball balance training on trunk control in individuals with sub-acute stroke

To assess the effect of conventional balance training on trunk control in individuals with sub-acute stroke

To compare the effect of Swiss ball balance training and conventional balance training on trunk control and mobility performance in individual with sub-acute stroke.

## Methods

### Study design

This study is a single center, randomized, crossover, assessor-blinded controlled trial involving individuals with a first-ever sub-acute stroke. Trained instructors will conduct the trials. Participants will be randomly assigned to one of two intervention sequences (AB or BA). The study will be conducted in the physiotherapy department at St Francis’ hospital. A trained researcher who is unaware of the participants’ group assignment will attend to the patients who would have obtain informed consent. Figure 1 shows the study flowchart.

**Figure 1.**
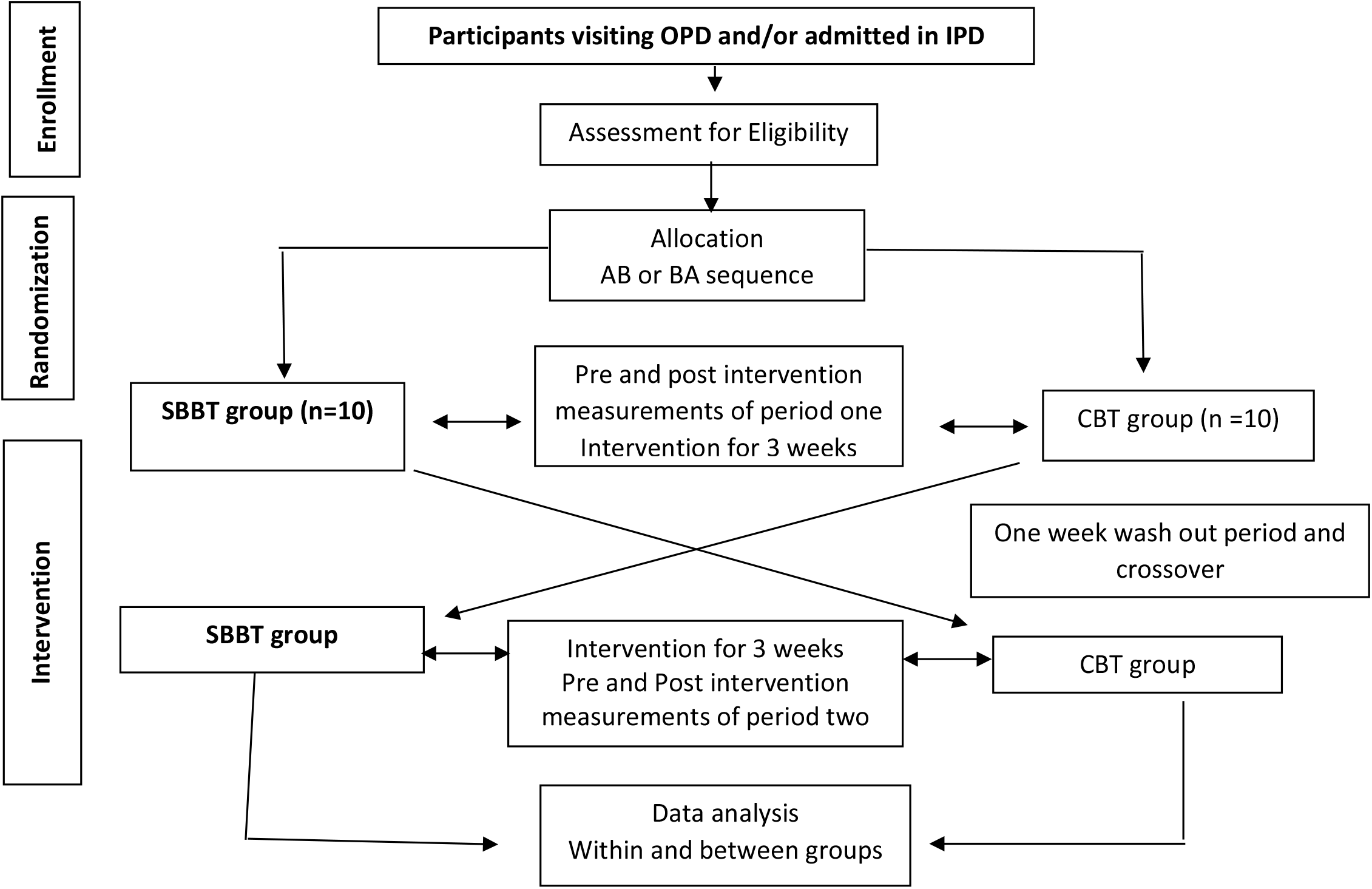
trial flow process.

### Participants

A total of 20 participants with sub-acute stroke fulfilling the eligibility criteria listed below will be included in the randomized controlled trial (RCT):

### Inclusion criteria

1. Stroke onset within 3 months prior to enrollment;
2. Willingness to participate in a 7 weeks study
3. Both right hemiplegic and left hemiplegic patients.
4. Male and female hemiplegic patients between the ages of 18 and 65 years
5. First-ever unilateral stroke

### Exclusion criteria

1. Cognitive impairment.
2. Uncontrolled hypertension (>180/120 mmHg)
3. Seizures.
4. Participation in other experimental rehabilitation studies.
5. Other reasons for exclusion (unable to mentally unable to participate, etc.)

### Recruitment procedure

Participants will be recruited from inpatient and outpatient departments, as well as through direct referrals to the physiotherapy department. Recruitment is scheduled to commence in October 2026. Eligible individuals will be invited to participate in the RCT. A trained research assistant will provide verbal information about the study in a private setting, either during or immediately following a routine care appointment. All research assistants will undergo training in a standardized recruitment protocol that incorporates evidence-based strategies to maximize enrollment and retention. Once a potential participant expresses verbal interest, written informed consent will be obtained by study personnel before formal enrollment. The overall trial flow is presented in Figure 1.

### Blinding and Randomization procedure

The principal investigator and all outcome assessors will remain blinded to group allocation. Following baseline assessment, participants who provide informed consent and meet eligibility criteria will be randomized in a 1:1 ratio. Allocation will be concealed using sealed, opaque envelopes prepared by an independent research assistant.

Treating physiotherapists will be aware of group assignments; however, outcome assessors and caregivers will remain blinded throughout the study. Endpoint assessments will be performed by two independent assessors, each with a minimum of two years of clinical experience in neurorehabilitation and standardized training in the administration of SBBT. To preserve blinding, assessors will not be involved in treatment delivery and will remain uninformed of group allocation

### Interventions

Participants randomized to either of the two treatment arms will receive the intervention in an AB/BA crossover format, consisting of SBBT and CBT. Following an initial 3-week intervention period and a one week washout period, participants will cross over to the alternate treatment arm, thereby receiving both interventions sequentially

## Intervention

### Swiss Ball Balance Training (SBBT)

SBBT will be delivered three times per week over a three-week period, with each session lasting approximately 45 minutes. The interventions will follow an adapted SBBT protocol from similar studies with exercise for individuals with sub-acute stroke (Shinde et al., 2023; Gokul et al., 2023).

Each session will be structured into five phases:

- Warm-up (8 minutes) – gentle preparatory movements to activate trunk and lower limb muscles.
- Balance in supine (5 minutes) – exercises performed lying on the ball to initiate core activation.
- Balance in prone (20 minutes) – progressive trunk and pelvic control activities emphasizing stability and coordination.
- Sitting exercises (20 minutes) – dynamic seated tasks targeting trunk control, balance, and functional mobility.
- Cool-down (7 minutes) – relaxation and stretching to promote recovery

### Conventional Balance training (CBT)

Participants randomized to the CBT arm will receive their usual treatment sessions, representing the standard care provided outside the study protocol. These sessions will include: Maintaining joint range of motion, practicing transfers between sitting and standing, Supporting seated posture (with or without assistance) and Performing coordination exercises

In addition, participants will engage in activities designed to improve balance and stability, thereby supporting overall functional mobility. Therapists will not be provided with a treatment guide, as the intervention reflects routine clinical practice. Conventional balance training will be delivered over a 3-week period.

### Data collection procedure and outcomes

Data will be collected using the Berg Balance Scale (BBS), the Trunk Impairment Scale (TIS), and the Timed Up and Go Test (TUG). Assessments will be conducted at baseline, at three weeks post-intervention, and three weeks following the second intervention period. These outcomes were selected to capture the anticipated effects of the intervention while also aligning with the majority of recommended core outcome measures in stroke rehabilitation trials (17).

An overview of the measures and outcomes is provided in Table 2

**Table 2.** Outcomes: T0, baseline; T1, evaluation at 3rd week and 7th week; BBS, Berg Balance Scale; TIS Trunk Impairment Scale; TUG, Timed Up and Go.

| Outcome domain | Measurement instrument | Swiss ball Balance Training |  | Conventional balance Training |  |
| --- | --- | --- | --- | --- | --- |
|  |  | T0 | T1 | T0 | T1 |
| Sociodemographic and descriptive data | Ad hoc questionnaire | x |  | x |  |
| Static balance | BBS | x | x | x | x |
| Dynamic balance and gait | TIS | x | x | x | x |
| Mobility | TUG | x | x | X | x |

### Participant characteristics

Participant characteristics such as Age, sex educational level, work status, marital status, smoking habits, address place of residence and if living alone or with others will be evaluated by a questionnaire.

### Outcomes

All participants will undergo assessments at three time points: baseline (A0), immediately following the first intervention period (A1), and after completion of the second intervention period (A2). Trained research assistants, who will receive standardized instruction and participate in consensus meetings, will conduct all evaluations to ensure methodological consistency and reliability. The primary outcome is the descriptive trunk balance status and the secondary outcome will be mobility as measured by the earlier stated tools.

### Berg Balance Scale (BBS)

The BBS is composed of 14 functional tasks, including reaching, standing on one leg, and controlled transfers. Each task is scored on a 0–4 scale, with a maximum of 56 points. Higher scores reflect better balance ability(18). 0–20 points indicates Severe balance impairment, 21–40 points indicates Moderate impairment and 41–56 points is Mild impairment (18).

The BBS demonstrates excellent reliability, with inter-rater reliability reported at approximately 0.97 (95% CI: 0.96–0.98) and intra-rater reliability at 0.98 (95% CI: 0.97–0.99). Absolute reliability varies, with minimal detectable change (MDC) estimates ranging between 2.8/56 and 6.6/56 points depending on the population studied(19).

### Timed up and Go (TUG)

The TUG test assesses dynamic balance and mobility. Participants are instructed to rise from a chair, walk three meters in a straight line, turn, return to the chair, and sit down, while the examiner records the time taken to complete the task. The TUG is widely used to estimate fall risk: scores below 10 seconds indicate minimal risk, scores between 10 and 20 seconds suggest moderate risk and increased vulnerability, and scores above 20 seconds reflect a high risk of falling (20). In individuals with stroke, the minimum detectable change (MDC) has been estimated at approximately 2.9 seconds, meaning improvements beyond this threshold are considered clinically meaningful (21). The sample size estimation was informed by previous studies employing the BBS and the TIS as primary outcome measures in stroke rehabilitation (22). Both instruments are validated and reliable tools that support clinical decision-making in this population (23). The calculation was based on the ability to detect a clinically meaningful difference in balance outcomes, as measured by the BBS and TIS.

In rehabilitation research, sample size determination is commonly guided by the minimal clinically important difference, defined as the smallest change in an outcome perceived as beneficial by patients and clinicians(24,25). To ensure both statistical power and clinical relevance, the required sample size was calculated as 10 participants per intervention arm (assuming a common standard deviation of 0.156, power = 80%, α = 0.05). Allowing for an anticipated 10% attrition rate, a total of 20 participants will be recruited.

#### 2.4 Study Sample

Participants will be recruited from the Physiotherapy Department at St. Francis Hospital. Patients diagnosed with stroke who attend the unit will be referred to a research assistant on the research team, who will screen them against the inclusion and exclusion criteria prior to inviting them to join the study. Eligible individuals will receive an information sheet and a verbal explanation of the study. They will be informed that they will be randomly assigned to one treatment arm (either the experimental or control group), later on they will switch the treatments. Those who agree to participate will sign an informed consent form, after which two research assistants from the research team will perform the baseline assessments

### Statistical analysis

Descriptive statistics will be reported for demographic data. Group comparisons between SBBT and CBT will be conducted at each evaluation time point using parametric tests (e.g., t-test) or non-parametric tests (e.g., Mann–Whitney U test, Wilcoxon test), as appropriate based on data distribution.

Between-group comparisons will be assessed across time points using either parametric or nonparametric methods, depending on the distributional characteristics of the data. In addition, longitudinal intragroup analyses will be performed to evaluate changes over time, employing parametric tests (e.g., paired t-test) or non-parametric alternatives (e.g., Wilcoxon signed-rank test), as appropriate.

All statistical analyses will be performed using Stata version 28.0 for Macintosh (Stata Corp LLC, College Station, TX, USA). Statistical significance will be set at p < 0.05. All randomized participants will be included in the intention-to-treat (ITT) analysis

### Dissemination and protocol amendments

The results, regardless of whether they are in favor of the intervention or comparator group, or inconclusive in relation to the study hypothesis, will be communicated in scientific papers, at relevant conferences. Any important protocol amendments will be reported to UNZASHREC and registered at ClinicalTrials.gov and presented in the primary RCT report.

### Discussion

This randomized crossover, assessor-blind controlled trial is the first to directly compare the effectiveness of SBBT with CBT in patients with sub-acute stroke at SFH. Stroke often results in impaired balance due to muscle weakness, atrophy, reduced muscle power, diminished cardiorespiratory fitness, and decreased functional performance, all of which contribute to gait disturbances and limitations in activities of daily living. The primary aim of this study is to evaluate the impact of SBBT versus CBT on static and dynamic balance, as well as mobility, in sub-acute stroke patients.

Targeted balance training is essential for restoring independence in functional tasks such as sit-to-stand transitions and turning movements, which are critical for daily living(26,27). Evidence suggests that interventions enhancing balance during the early recovery phase may also facilitate neuroplasticity and functional reorganization of the nervous system (28).

Studies have demonstrated that SBBT improves postural control and functional outcomes in stroke rehabilitation (28). The dynamic and unstable surface of the Swiss ball challenges trunk stability, thereby promoting activation of core musculature and enhancing proprioceptive feedback(7,29). Moreover, rhythmic bouncing and rocking movements on the Swiss ball have been shown to increase alertness by stimulating the vestibular system and activating the reticular formation (2). In contrast, CBT typically emphasizes exercises performed on stable surfaces, which may improve strength but provide less stimulation for dynamic postural adjustments (16). Consequently, incorporating Swiss ball exercises may accelerate improvements in trunk control, balance confidence, and functional mobility compared to conventional methods (30).

Literature has reported that Swiss ball training significantly enhances core strength and proprioception, as the unstable surface necessitates continuous neuromuscular engagement (30, 16). Similarly, Muniyar et al. found that trunk exercises performed on a Swiss ball were more effective than those performed on a plinth in improving trunk control and functional balance in stroke patients, though their study focused on acute rather than sub-acute cases(22). This distinction will be important for contextualizing the findings from the present study. Furthermore, Swiss ball training has the potential to improve muscle strength, cardiorespiratory fitness, and performance in activities of daily living, while simultaneously reducing the risk of secondary complications associated with inactivity (31). By contrast, CBT, while efficacious, is often associated with a delayed therapeutic response, potentially prolonging inactivity and thereby increasing the risk of disability (32).

In summary, SBBT may offer distinct advantages over CBT in sub-acute stroke rehabilitation by promoting dynamic trunk control, enhancing proprioceptive feedback, and improving functional mobility (7,33). The findings of this study could inform clinical practice and contribute to the development of accessible, effective rehabilitation strategies for patients recovering from stroke.

## Conclusion

This study will provide preliminary evidence on the comparative effectiveness of SBBT and CBT in improving trunk control, balance, and mobility among individuals with sub-acute stroke. Findings are expected to demonstrate that SBBT produces greater gains in trunk stability, balance performance, and functional mobility compared to CBT. These results will support the design of context-specific rehabilitation programs and may inform the development of accessible, costeffective strategies, particularly for low-resource settings. Moreover, the study can serve as a foundation and motivation for future research, encouraging scholars to conduct larger, more representative studies to strengthen the evidence base.

### Ethical Approval of the study

Ethical approval for this study was obtained from the University of Zambia, School of Health Sciences Research Ethics Committee (Protocol ID: 2023270588). Additional authorization to conduct the research was granted by the National Health Research Authority (REF: NHRA-3832/10/07/2026) and further authority will be sought from St. Francis Hospital. The trial has been registered with the Pan African Clinical Trials Registry (PACTR202605730696438). The principal investigator has also completed formal training in Good Clinical Practice.

All participants will be provided with an information sheet outlining the purpose and nature of the study, the data collection procedures, and their rights as participants. This will ensure that they are able to make an informed and voluntary decision regarding participation. Participants will be reminded of their right to withdraw from the study at any time without the need to provide justification. Confidentiality and anonymity will be strictly upheld; no participant names will appear on questionnaires or study records.

## Data Availability

No datasets have generated or analysed yet during the current study. However, all relevant data from this study will be made available upon completion

## Acknowledgements

The authors gratefully acknowledge the participants who will take part in this trial. We hope to extend our sincere appreciation to the administration and staff of St. Francis Hospital for the anticipated support and cooperation. We are also profoundly grateful to Dr. Miriam Mapulanga for her invaluable mentorship and guidance throughout the development of this protocol

## Declaration of conflicting interests

The authors declare no conflicts of interest regarding the publication of this paper

## Funding

This study was not funded

## Notes

### Competing Interest Statement

The authors have declared no competing interest.

### Clinical Trial

PACTR20265730696438

### Author Declarations

UNIVERSITY OF ZAMBIA HEALTH SCIENCES RESEARCH ETHICS COMMITTEE P. O. Box 50110 Tel: +260966435366 Lusaka, 15101 Tel: +260977445813 Zambia IRB no: 00011000

## References

1. Awais D, Batool S, Ahmad A, Ali Aftab A, Naqvi R. Comparison of Routine Physical Therapy with And Without Core-Stability Exercises on Dynamic Sitting Balance and Trunk Control in Sub-Acute Ischemic Stroke Patients: Routine Physical Therapy with and without Core-Stability Exercises. Pak Biomed J. 2022 Sep 30;31–5. doi:10.54393/pbmj.v5i9.796

2. Shinde GM, Shinde DDP, Meshram DVK. Effect of Swiss ball training versus conventional physiotherapy in static, dynamic balance and postural stability using x-16 scale in hemiplegic patients: a comparative study. vol. 13. 2023; 13(2).

3. Liu H, Yin H, Yi Y, Liu C, Li C. Effects of different rehabilitation training on balance function in stroke patients: a systematic review and network meta-analysis. Arch Med Sci. 2023 Jun 21. doi:10.5114/aoms/167385

4. Khurana Y, Devi M, Kaur A, Subramanian T, Mani S. Is Swiss-ball-based exercise superior to plinth-based exercise in improving trunk motor control and balance in subjects with sub-acute stroke? A pilot randomized control trial. Physiother Q. 2022 Sep 26; 30(3):72–8. doi:10.5114/pq.2021.103558

5. Agina Widyaswara Suwaryo P, Santoso EB, Utoyo B. The Effectiveness of Swiss Ball Exercise to Increase Balance and Mobility of Patient with Stroke. Babali Nurs Res. 2023 Apr 30;4(2):185–94. doi:10.37363/bnr.2023.42135

6. Feigin VL, Abate MD, Abate YH, Abd ElHafeez S, Abd-Allah F, Abdelalim A, et al. Global, regional, and national burden of stroke and its risk factors, 1990–2021: a systematic analysis for the Global Burden of Disease Study 2021. Lancet Neurol. 2024 Oct; 23(10):973–1003. doi:10.1016/S1474-4422(24)00369-7

7. Khurana Y, Devi M, Kaur A, Subramanian T, Mani S. Is Swiss-ball-based exercise superior to plinth-based exercise in improving trunk motor control and balance in subjects with sub-acute stroke? A pilot randomized control trial. Physiother Q. 2022 Sep 26; 30(3):72–8. doi:10.5114/pq.2021.103558

8. Ishiwatari M, Honaga K, Tanuma A, Takakura T, Hatori K, Kurosu A, et al. Trunk Impairment as a Predictor of Activities of Daily Living in Acute Stroke. Front Neurol. 2021 Jun 17; 12:665592. doi:10.3389/fneur.2021.665592

9. Khallaf ME. Effect of Task-Specific Training on Trunk Control and Balance in Patients with Subacute Stroke. Neurol Res Int. 2020; 2020:5090193. doi:10.1155/2020/5090193 PubMed PMID: 33294224; PubMed Central PMCID: PMC7688364.

10. Gokul M, Dhasaradharaman K, Robert F. A Comparative Study on the Swiss Ball Training and Conventional Balance Training Versus Proprioceptive Neuromuscular Facilitation Pattern for Improving Trunk and Balance Control in Sub-Acute Stroke. Int J Health Sci Res. 2023 Dec 5;13(12):58–63. doi:10.52403/ijhsr.20231207

11. Kim YW, Yoon SY. The Safety and Efficacy of Balance Training on Stroke Patients with Reduced Balance Ability: A Meta-Analysis of Randomized Controlled Trials. Brain Neurorehabilitation. 2024;17(3):e15. doi:10.12786/bn.2024.17.e15

12. Alagappan P, Arumugam M, Veyilmuthu R. Efficacy of Swiss Ball Therapy in Balance Rehabilitation of Hemiplegic Stroke Participants. 2019;(6).

13. Noreen A, Lu J, Xu X, Jiang H, Hua Y, Shi X, et al. comparing the effects of Swiss-ball training and virtual reality training on balance, mobility, and cortical activation in individuals with chronic stroke: study protocol for a multi-center randomized controlled trial. Trials. 2024 Oct 14;25(1):677. doi:10.1186/s13063-024-08532-9

14. Ravichandran H, Sharma HR, Haile TG, Gelaw AY, Gebremeskel BF, Janakiraman B. Effects of trunk exercise with physioball to improve trunk balance among subjects with stroke: a systematic review and meta-analysis. J Exerc Rehabil. 2020 Aug 25; 16(4):313–24. doi:10.12965/jer.2040292.146

15. Li X, He Y, Wang D, Rezaei MJ. Stroke rehabilitation: from diagnosis to therapy. Front Neurol. 2024 Aug 13;15:1402729. doi:10.3389/fneur.2024.1402729

16. Saraiva J, Rosa G, Fernandes S, Fernandes JB. Current Trends in Balance Rehabilitation for Stroke Survivors: A Scoping Review of Experimental Studies. Int J Environ Res Public Health. 2023 Sep 26;20(19):6829. doi:10.3390/ijerph20196829

17. Van Criekinge T, Heremans C, Burridge J, Deutsch JE, Hammerbeck U, Hollands K, et al. Standardized measurement of balance and mobility post-stroke: Consensus-based core recommendations from the third Stroke Recovery and Rehabilitation Roundtable. Neurorehabil Neural Repair. 2024 Jan;38(1):41–51. doi:10.1177/15459683231209154

18. Önal B, Köse N, Önal ŞN, Zengin HY. Validity and Reliability of the Berg Balance Scale in Different Tele-Assessment Methods in Patients With Stroke. J Eval Clin Pract. 2025 Jun;31(4):e70141. doi:10.1111/jep.70141

19. Matsumoto D, Fujita T, Kasahara R, Tsuchiya K, Iokawa K. Screening cutoff values to identify the risk of falls after stroke: A scoping review. J Rehabil Med. 2024 Oct 24;56:jrm40560. doi:10.2340/jrm.v56.40560

20. Reis M, Teixeira M, Carvão C, Martins AC. Validity and Reliability of the Self-Administered Timed Up and Go Test in Assessing Fall Risk in Community-Dwelling Older Adults. Geriatrics. 2025 Apr 29;10(3):62. doi:10.3390/geriatrics10030062

21. Kim MK. The effects of trunk stabilization exercise using a Swiss ball in the absence of visual stimulus on balance in the elderly. J Phys Ther Sci. 2016 Jul;28(7):2144–7. doi:10.1589/jpts.28.2144 PubMed PMID: 27512284; PubMed Central PMCID: PMC4968524.

22. Shinde SG, Bafna PS. Comparison of Swiss ball exercises versus conventional therapy on improving trunk control in patients with acute and subacute stroke. Int J Health Geogr. 2019 Mar;4(1):385

23. Mapulanga, M. (2015). Exploration and determination of the process of care of stroke in Zambia (Master’s thesis, University of the Western Cape). University of the Western Cape.

24. Jaeschke R, Singer J, Guyatt GH. Measurement of health status. Control Clin Trials. 1989 Dec;10(4):407–15. doi:10.1016/0197-2456(89)90005-6

25. Revicki D, Hays RD, Cella D, Sloan J. Recommended methods for determining responsiveness and minimally important differences for patient-reported outcomes. J Clin Epidemiol. 2008 Feb;61(2):102–9. doi:10.1016/j.jclinepi.2007.03.012

26. Du M, Chen L, Xia L, Li Y, Ma E, Hu Z, et al. Effectiveness of different exercise interventions on balance and cognitive functions in stroke patients: A network meta-analysis. BMC Sports Sci Med Rehabil. 2025 Aug 27;17(1):250. doi:10.1186/s13102-025-01267-3 PubMed PMID: 40866917; PubMed Central PMCID: PMC12382136.

27. Chen X, Gan Z, Tian W, Lv Y. Effects of rehabilitation training of core muscle stability on stroke patients with hemiplegia: Rehabilitation training of stroke patients. Pak J Med Sci. 2020 Mar 4;36(3). doi:10.12669/pjms.36.3.1466

28. Suwaryo PAW. Swiss ball exercise post-stroke with hemiparesis to improve mobility: a randomised controlled trial.

29. A C, S G. The Effect of Swiss Ball Exercises Versus Plinth Exercises on Trunk Control and Balance among Post-Stroke Patients. Int J Sci Res IJSR. 2025 Dec 11;530–4. doi:10.21275/SR251207004357

30. Pandya RH, Sutaria JM. Effects of Swiss Ball Trunk Exercises on Trunk Control and Functional Balance in Post Stroke Patients - An Interventional Study. 2021;(1).

31. Mishra DrMK, Kanaujia DrS. Effect of swiss ball exercises on muscular strength and endurance in male students. Int J Phys Educ Sports Health. 2021 Nov 1;8(6):90–3. doi:10.22271/kheljournal.2021.v8.i6b.2290

32. Unger J, Chan K, Lee JW, Craven BC, Mansfield A, Alavinia M, et al. The Effect of Perturbation-Based Balance Training and Conventional Intensive Balance Training on Reactive Stepping Ability in Individuals With Incomplete Spinal Cord Injury or Disease: A Randomized Clinical Trial. Front Neurol. 2021 Feb 2;12:620367. doi:10.3389/fneur.2021.620367

33. Noreen A, Lu J, Xu X, Jiang H, Hua Y, Shi X, et al. Comparing the effects of Swiss-ball training and virtual reality training on balance, mobility, and cortical activation in individuals with chronic stroke: study protocol for a multi-center randomized controlled trial. Trials. 2024 Oct 14;25(1):677. doi:10.1186/s13063-024-08532-9

